# Understanding the pathogenesis of uveitis in Ebola virus disease survivors: a study protocol for clinical, molecular virologic, and immunologic characterization

**DOI:** 10.1101/2025.05.19.25327799

**Authors:** Caleb D. Hartley, Susanne Linderman, Tolulope Fashina, Laura Ward, Carolyn Drews-Botsch, Catherine Pratt, Sanjana Kuthyar, Alcides Filho Fernandes, Ye Huang, Charlene Choo, Nam Nguyen, Jessica Carag, Jill Morgan, Colleen S. Kraft, Angela Hewlett, David Brett-Major, John S. Schieffelin, Robert F. Garry, Donald Grant, Grant A. Justin, Christopher D. Conrady, Justine R. Smith, Brent R. Hayek, Shiama Balendra, Nisha Acharya, Thuy Doan, Anais Legand, Pierre Formenty, Xiankun Zeng, Ibrahim Conteh, Matthew J. Vandy, Lloyd Harrison-Williams, Jalikatu Mustapha, Zikan Koroma, Michael Wiley, Ian Crozier, Jean-Claude Mwanza, Jessica G. Shantha, Rafi Ahmed, Steven Yeh, SMILE and EVICT-VR Investigator Study Groups

## Abstract

The 2013-2016 Western African outbreak of the Ebola virus disease (EVD), the largest recorded outbreak since the discovery of Ebola virus (EBOV) in 1976, destabilized local health systems and left thousands of survivors at risk for post-acute sequelae, including vision-threatening uveitis. In an EVD survivor with severe panuveitis, the detection of persistent EBOV in the aqueous humor, long after clearance of acute viremia, focused clinical and research attention on the host-EBOV interaction in the unique terrain of ocular immune-privilege. Despite the recognition that uveitis is common and consequential in EVD survivors, our understanding of pathogenesis is extremely limited, including the contributions of viral persistence and ocular-specific and systemic immune responses to disease expression.

In this study protocol, we outline a multifaceted approach to characterize EVD-associated intraocular inflammation (EVD-IOI), including the clinical phenotype and complications; the presence of EBOV (or EBOV RNA/antigen) in ocular fluids and tissues; and associated local ocular-specific and peripheral immune responses. We utilize an observational cohort design, which includes EVD survivors and close contacts of EVD survivors (i.e., no documented history of EVD), and we propose disease (clinical examination and imaging), as well as molecular, virologic and immunologic characterization, to meet research objectives. Comprehensive findings emerging from the research will inform local stakeholders and global partners to understand and effectively address the individual and public health implications of EVD-associated uveitis, including to optimize clinical decision-making and medical intervention, identify potential ocular and peripheral biomarkers of viral persistence and ocular disease, and ensure effective infection prevention and control.

## INTRODUCTION

Since the Ebola virus (EBOV) was first identified in 1976 as the cause of Ebola virus disease (EVD) in the Democratic Republic of the Congo (DRC, formerly Zaire), more than 40 outbreaks of disease caused by orthoebolaviruses have been reported to date [1–4]. Outbreaks have varied in magnitude and duration but, regardless of size, have been devastating to affected individuals and communities. Past outbreaks have also presented medical, scientific, and logistical challenges to the global health community, given the need to improve individual and public health outcomes both for acute EVD and for post-acute sequelae in EVD survivors.[5]

Prior studies from Western Africa and Central Africa have described post-acute sequelae in EVD survivors, including eye disease, joint disease, mental health disorders, and the risk of EBOV persistence in immune privileged tissues or fluids of the eye [6], central nervous system [7], and reproductive organs [8]. Despite a recent increase in our understanding, considerable uncertainty about the individual and public health consequences of post-acute sequelae in EVD survivors warrants further research to inform preparedness for future events; recent or ongoing filovirus disease (FVD) outbreaks in Central [9, 10] and Western Africa [11] highlight the importance of this research, not just for EVD, but for FVD writ large.

Uveitis, a sight-threatening inflammatory eye disease, is the most common ocular complication associated with EVD. Though first described in four survivors of the DRC Kikwit (1995-1996) outbreak [12], EVD-associated uveitis first received significant clinical and research attention during and after the historically largest Western African (2013-2016) EVD outbreak [5, 13–15]. Notably, in a repatriated United States healthcare worker and EVD survivor who developed severe, sight-threatening panuveitis during convalescence, infectious EBOV was detected in the aqueous humor (AH) long after clearance of acute viremia [5], calling attention to the role of viral persistence in vision-threatening inflammation in the immune-privileged ocular tissues of EVD survivors.

Data from multiple observational cohorts in Western Africa suggest a surprisingly high (13%-34%) prevalence of uveitis in EVD survivors [13, 14, 16, 17] and the need to understand the clinical manifestations and complications in order to optimize EVD survivor eye care. At baseline enrollment, the NIH PREVAIL Study described a higher (26% versus 12%) prevalence of uveitis in Liberian EVD survivors as compared to close-contact controls, respectively; notably, the prevalence of uveitis (active or inactive) in survivors continued to increase over their first year of follow-up, suggesting incident disease [18]. In another retrospective study from Liberia in collaboration with ELWA Hospital, Shantha et al reported that 20-25% of 100 examined EVD survivors had clinical evidence of prior or current uveitis, and nearly 40% of survivors with uveitis developed blindness in the affected eye [14]. Eghrari et al described additional post-uveitic complications in EVD survivors, including lower intraocular pressure, posterior synechiae, cataract development, and retinal scarring [19].

As evidence mounted that visually-significant post-uveitic cataract development was a common and potentially surgically addressable complication, uncertainty about the potential for EBOV to persist in the intraocular fluids and tissues of EVD survivors with cataract required urgent clinical and research attention. The Ebola Virus Persistence in Ocular Tissues and Fluids (EVICT) Study, conducted in 2016-2017 in Sierra Leone [20], evaluated sampled AH for EBOV RNA by reverse transcription-quantitative polymerase chain reaction (RT-qPCR) in 50 EVD survivors, with the majority undergoing an assessment prior to cataract surgery.

In the EVICT study, and in subsequent similar investigation in Liberian EVD survivors [15], EBOV RNA was not detected in the AH of EVD survivors with cataract, though it should be noted that ocular fluid sampling generally occurred long after primary uveitis episodes and in eyes without active inflammation. In a non-human primate survivor rescued by an EBOV-specific monoclonal antibody therapeutic, clinical uveitis with destructive ocular immunopathology has been associated with detection of EBOV RNA and high levels of EBOV-specific immunoglobulin G (IgG) in the vitreous humor > 3 months after acute infection; retrospective analyses of archived tissues of treated NHP survivors have also identified EBOV RNA in macrophages at the vitreo-retinal interface [21, 22].

Given these observations, as well as the finding of the late elevation of EBOV antigen-specific IgG4 in association with isolated cases of EBOV persistence [23], we hypothesized that antigen-specific immune responses associated with ocular persistence of EBOV, specifically within the vitreous humor, might provide a relevant biomarker and determine the pathophysiology of uveitis in Western African EVD survivors.

By leveraging the expertise of a uniquely qualified collaborative group of investigators, we aim to characterize the prevalence, clinical, virologic, and immunologic features of EVD-associated uveitis in EVD survivors and appropriate control populations. These assessments include a combination of rigorous clinical ophthalmic evaluation, assessment of the vitreous for EBOV RNA persistence, and characterization of systemic and ocular EBOV antigen-specific immune responses.

The specific aims of this research initiative include the following:

1. Determine the prevalence, ophthalmic disease spectrum, functional impact, and risk factors for uveitis in EVD survivors as compared to household members with close contact with the patient during their acute illness.
2. Determine whether EBOV antigen or nucleic acid can be detected in vitreous fluid by molecular and immunologic assessment of vitreous fluid (including RT-PCR testing, in-situ hybridization (ISH), or immunohistochemical (IHC) approaches)
3. Characterize EBOV-specific antibody and cell-mediated responses in the peripheral blood and ocular fluid of EVD survivors with uveitis.

The study findings will further define the clinical spectrum of uveitis associated with EVD, improve our understanding of the potential for EBOV to persist in intraocular reservoirs, and potentially identify novel immunologic biomarkers of intra-ocular viral persistence. Knowledge generated may inform strategies to detect, prevent, and mitigate EVD-associated uveitis in the future.

## METHODS

### Study Design and Setting

The clinical research program includes three studies developed around each research aim to inform management approaches to eye disease in EVD survivors (Figure 1). Each study will be carried out at the Lowell and Ruth Gess Eye Hospital (LRGEH) in Sierra Leone and Connaught Government Hospital Eye Clinic, alongside in-country collaborators from the Sierra Leone Ministry of Health (MOH) National Eye Programme.

**Figure 1.**
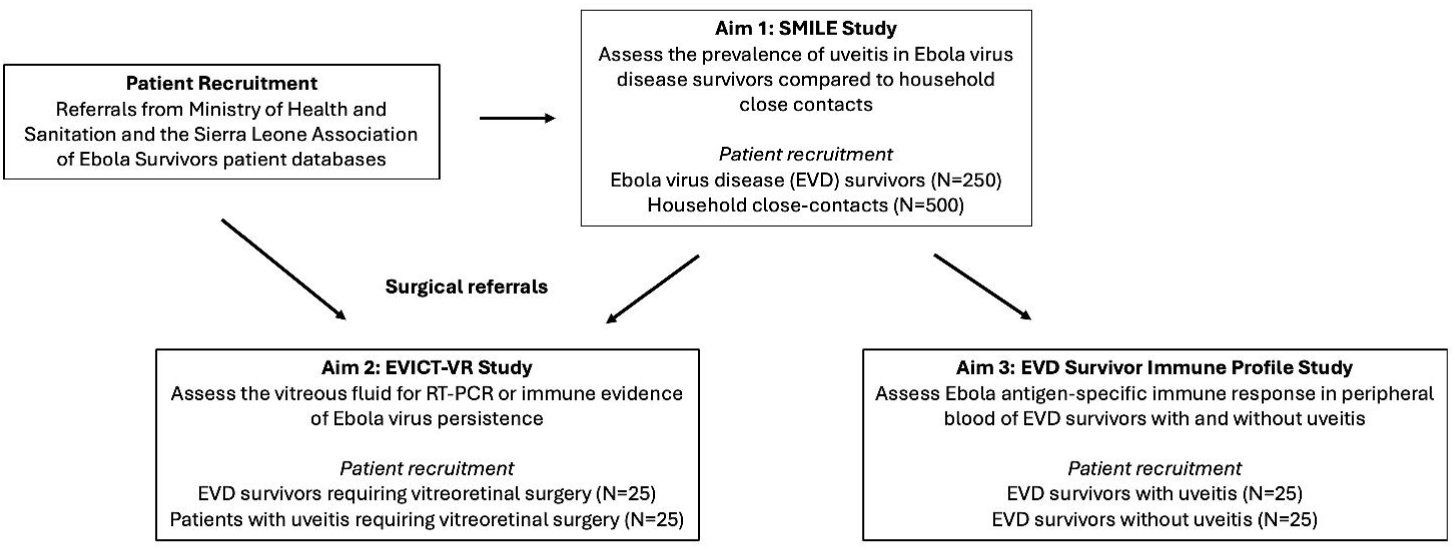
Study schematic highlighting the three studies that will address research aims. Aim 1 will assess EVD survivors for the prevalence of uveitis and ophthalmic outcomes and is entitled the Sierra Leone Ministry of Health and Lowell and Ruth Gess Hospital Ebola Survivor (SMILE) study, in which at least 750 patients will be recruited (i.e., 250 EVD survivors and 500 close-contacts). In Aim 2, the Ebola Virus Persistence in Ocular Tissues and Fluids Study-Vitreoretinal (EVICT-VR) will assess vitreous fluid for molecular and immunologic evidence of EBOV antigen or nucleic acid persistence. In Aim 3, EBOV antigen-specific immune responses will be assessed in EVD survivors with and without uveitis. The referral process is outlined by the arrows and include Ministry of Health and Sanitation, Sierra Leone engagement, along with the Sierra Leone Association of Ebola Survivors.

The first observational cohort study, termed the **S**ierra Leone **Mi**nistry of Health and **L**owell and Ruth Gess Eye Hospital **E**bola Survivor ***(SMILE)*** Study will assess the prevalence of uveitis following acute infection, ocular complications secondary to uveitis (e.g., posterior synechiae, cataract, cystoid macular edema) and visual acuity impact in EVD survivors compared to a group of close contact controls without a history of EVD.

A second, cross-sectional study, termed the **E**bola **Vi**rus Persistence in O**c**ular **T**issues and Fluids Study – **V**itreo**r**etinal Surgery ***(EVICT-VR)***, which will focus on EVD survivors with indications for vitreoretinal surgery. Molecular virologic and immunologic characteristics (in vitreous and peripheral blood) will be investigated in EVD survivors undergoing vitreoretinal surgery (for diagnostic or therapeutic indications) and compared to patients requiring vitreoretinal surgery with a uveitis syndrome not known to be associated with EVD.

A third cross-sectional study will assess the peripheral blood B- and T-cell and Ebola antigen-specific antibody responses of EVD survivors with and without uveitis.

### Collaboration and Roles

This work integrates a multidisciplinary team including expertise from a range of specialties including ophthalmology, infectious diseases, epidemiology, genomics, and immunology to meet the aims of the project. Longstanding partnerships with the Sierra Leone MOH and the Sierra Leone Association of Ebola Survivors (SLAES) will support patient recruitment.

### Ethics/Institutional Review Board

Institutional Review Board (IRB) approvals were obtained from the University of Nebraska Medical Center (IRB approval number 0307-21-FB) and Emory University School of Medicine (IRB00108429). Research Ethics Board approval was obtained from the Ministry of Health Office of Ethics Approval in Sierra Leone.

### Patient and Public Involvement

Patients or the public were not involved in the design, conduct, reporting, or dissemination plans of our research.

### Patient Recruitment Strategy for Research Aims

#### Aim 1

Using a conservative prevalence estimate of 10% of uveitis in EVD survivors and compared to estimate of 1% in controls, a sample size of 200 per group is needed to achieve a 95% confidence level and greater than 80% power of detecting a significant difference in uveitis prevalence between groups. EVD survivors and close-contacts will be recruited in a 1:2 ratio for a minimum survivor recruitment of 250 patients and 500 comparison subjects. Recruitment of EVD survivors will be continued beyond this goal where possible to assess risk factors for uveitis development within EVD survivor populations.

#### Aim 2

Fifty individuals will be recruited from SLAES and MOH screening and referral processes. These include 25 EVD survivors with or without uveitis who require vitreoretinal surgery and 25 patients with uveitis but without a history of EVD who require vitreoretinal surgery.

#### Aim 3

Fifty individuals will be assessed from EVD survivors referred from the MOH and SLAES, some of whom may have been evaluated in the context of Aim 1. This aim will assess individual immunologic responses and will include 25 EVD survivors with uveitis and 25 EVD survivors without uveitis.

### Participant Characteristics

#### Inclusion/Exclusion Criteria

##### Aim 1

Identification of EVD survivors and their first-degree close contacts (e.g., individuals who lived in the same place of residence and were not known to have developed EVD) will be facilitated by referral from MOH and SLAES. Those persons with positive serum EBOV IgG and/or original positive EBOV RT-qPCR results from MOH databases will be considered to be EVD survivors. Household close contacts of EVD survivors will also undergo serologic testing for anti-EBOV-X IgG; those considered anti-EBOV IgG-positive will be included as EVD survivors but analyzed as a separate group. Survivors of EVD and seronegative household close contacts (without history of EBOV RT-qPCR positivity) will be recruited in a 1:2 ratio. Exclusion criteria include evidence of active EVD or acute hemorrhagic fever, as well as the inability to complete initial or follow-up eye examinations, and the inability to provide consent.

##### Aim 2

Fifty patients with vitreoretinal pathology (e.g., epiretinal membrane, vitreomacular traction, tractional retinal detachment, vitreous opacities) that warrants surgery for diagnostic and therapeutic indications will be recruited from SLAES and MOH referrals. EVD survivors with and without uveitis (n=25) will comprise the EVD survivior cohort. Inclusion criteria for the non-EVD survivor cohort is patients with uveitis and no history of EVD and negative for anti-EBOV IgG antibodies (n=25). Exclusion criteria include those persons who are deemed unlikely to have a beneficial improvement in visual acuity, along with those unable to undergo surgery for medical or other reasons.

##### Aim 3

In order to assess the immunologic responses in the peripheral blood of EVD survivors, 25 EVD survivors with uveitis and 25 EVD survivors without uveitis will be recruited from SLAES and MOH referrals.

#### Patient Recruitment

The broad referral base for patient recruitment and community mobilization arises from the longstanding collaboration of our team with MOH and partnering organizations such as SLAES. Our team has partnered with MOH to coordinate care, research, and educational activities in Sierra Leone. We also previously partnered with MOH, Partners in Health, and United States Agency for International Development (USAID) affiliates for ophthalmic screening EVD survivors, and eventually established a clinical registry of data collected from these survivors. In all, these partnerships will enable us to successfully recruit participants during the designated study period.

#### Informed Consent

Written informed consent will be obtained for all patients, and assent and consent by a legal guardian as appropriate. Consent will be obtained by the physician performing the procedures, with translators present should a language barrier exist. Refusal of participation will not impact current or future care.

Individuals who meet eligibility criteria will be given the opportunity to ask questions and privately discuss with their family members prior to signing the informed consent documentation. Questions will be answered as they are asked. Individuals who provide informed consent can withdraw at any time without impact on their care. No identifying patient data will be reported in manuscripts or other research outputs.

### Status and Timeline of Study

Recruitment of participants into the study began on 16/03/2022 and is currently ongoing for both the SMILE and EVICT-VR arms. Participant recruitment is expected to be completed by 31/07/ 2025. Data collection to assess peripheral blood B- and T-cell profiles, as well as Ebola antigen-specific antibody responses in EVD survivors, is also ongoing, with anticipated completion by 31/03/2026.

### Study Procedures

#### Aim 1

Aim 1 entails clinical ophthalmic evaluation and peripheral blood sampling to assess for anti-EBOV IgG to sort by serologic status. The comprehensive clinical evaluation includes collection of historical and demographic information, as well as visual acuity assessment, intraocular pressure measurement, ophthalmic examination by a study clinician-investigator, color fundus photography, and optical coherence tomography (OCT).

Historical and demographic information to be collected includes age, sex, EVD treatment unit (ETU) setting, date of EVD diagnosis, systemic symptoms at the time of acute EVD, and pertinent ophthalmic history such as uveitis prior to EVD, ophthalmic therapies received, and any available MOH eye examination data. In addition, EBOV RT-qPCR cycling threshold data will be captured from MOH databases where available.

The case definition for *EVD-associated uveitis* employed in this study is as follows: Evidence of current or prior uveitis and/or its sequelae with either vision loss or symptoms that occurred during or after EBOV infection and were not related to trauma, prior illness or infection, or congenital disease. Clinical features that will be included for a case definition of uveitis include the presence of the following: keratic precipitates, posterior synechiae (PS), uveitic cataract (dense white cataract with PS), chorioretinal scarring, and/or band keratopathy if other signs of uveitis are present. Uveitis will be classified as active or inactive, predicated on the presence of anterior chamber (AC) cell, AC flare and/or vitreous haze (VH), as outlined in the Standardization of Uveitis Nomenclature (SUN) guidelines [24]. Patients with evidence of uveitis and/or its sequelae will have a laboratory-based work-up for infections associated with uveitis that may include serologic testing for toxoplasmosis, tuberculosis, syphilis, and human immunodeficiency virus (HIV) infection if there are risk factors, systemic findings, or ophthalmic exam features suggestive of these conditions.

Ophthalmic assessment by a study clinician will include the best-corrected visual acuity with an Early Treatment of Diabetic Retinopathy Study (ETDRS) chart at 4 meters, and intraocular pressure will be measured with a Tono-Pen Avia ® Tonometer (Reichert, Depew, NY). Tumbling “E” at 4 meters will be utilized for patients where a language barrier exists, or the patient is unable to read characters on the EDTRS chart.

Slit lamp and funduscopic examination will be used to define the anatomic location and extent of uveitis (anterior, intermediate, posterior, and pan-uveitis), along with structural complications associated with uveitis. Anterior chamber cell, AC flare, and VH scores will be documented according to SUN guidelines [24]. Slit lamp photographs will be captured with an iPhone at 10X magnification, and color fundus photography will be performed using a specialized fundus camera (Clarus 700, Carl Zeiss Meditec, Dublin, CA, USA).

Color fundus photography, OCT, and FFA, where indicated, will be used to characterize ultrastructural complications associated with uveitis. The OCT device (Cirrus® 5000 HD-OCT, Carl Zeiss Meditec, Dublin, CA, USA) will capture high-resolution images of complications in the vitreous and retina, retinal edema, choroidal thickening, and epiretinal membrane formation. Fundus angiographic images will specifically capture irregularities of the choroidal and retinal vasculature, to be correlated with other clinical findings. B-scan ultrasonography will be performed to view the posterior segment of the eye when media opacity prevents clear visualization of the retina and vitreous.

#### Aim 2

Patients recruited for Aim 2 will undergo pars plana vitrectomy (PPV) surgery for detailed evaluation of ocular fluid and tissue with infection prevention and control (IPC) precautionary measures outlined in the Standard Operating Procedure (SOP) – Vitreoretinal Surgery (Supplemental Material). During PPV surgery, an undiluted vitreous specimen of 1.0-1.5 mL will be collected for detection of EBOV RNA by RT-qPCR, in addition to EBOV-antigen-specific IgG and total IgG from paired vitreous and serum samples for calculation of the Goldmann-Witmer Coefficient (GWC). Laboratory technicians will be masked as to the group status of patient samples.

#### Aim 3

In Aim 3, peripheral blood of 25 EVD survivors with uveitis and 25 EVD survivors without uveitis will be sampled for analysis, along with the ocular fluid of 25 EVD survivors with uveitis. Peripheral phenotyping of B-cell and T-cell will occur as follows: peripheral blood will be centrifuged to isolate peripheral blood mononuclear cells, either in cell preparation tubes or on a Histopaque gradient. T-cell populations will be identified and activation status will be characterized via flow cytometry. EBOV antigen-specificity will be delineated using EBOV-antigen tetramers or intracellular cytokine staining following peptide stimulation.

To amplify sequences of lymphocyte receptors, RT-qPCR will be performed on undiluted vitreous samples. These sequences will be compared to previously defined EBOV-specific B-cell and T-cell clonotypes. They will also be compared to sequences from sorted tetramer binding T-cells or GP-binding B-cells from the peripheral blood to elucidate if EBOV-specific clonotypes are present within the eye.

### Data Management

Case report forms (CRFs) will be created using Adobe InDesign and approved by a clinician, a data manager, and a biostatistician. The CRFs will be programmed to DFNet (Seattle, Washington) and completed as paper forms on site at the time of the visit as study subjects are examined. Study personnel will quality check the CRF for completeness, correctness, and absence of identifying information prior to the subject leaving the study site. All CRFs will be stored locked in a patient-designated binder, and pdf-format files of the CRFs will be uploaded to the database *Pathogenesis of Uveitis in Ebola Survivors* using DFsend. This will sort the CRFs into the correct study, visit, and plate (a DataFax object corresponding to a unique CRF page) based on the identification barcode at the top of each CRF. Those lacking barcodes will be stored in a router, which a human validator will check twice weekly for sorting into the appropriate study, visit, and plate.

Once uploaded into DFcollect, DFnet will collate the CRF data into the appropriate data field. The human validator will quality check for accuracy and completeness of the forms in the system. Problems will be queried with the onsite study team in a weekly report. All validated electronic data will then be stored in the *Pathogenesis of Uveitis in Ebola Survivors* database and marked as validated. Edit checks will be programmed into the database, and discrepancies will be flagged for review automatically. A quality control report with all compiled edit checks will be created monthly. This report will not contain identifying information, and site researchers will have the opportunity to correct or comment on each discrepancy and edit check. As needed, corrections will be made on the paper CRFs by the site researchers and rescanned for data corrections.

All protected health information collected as part of this study will be de-identified according to established IRB and Research Ethics Board guidelines, and kept in password-protected, encrypted files. The database *Pathogenesis of Uveitis in Ebola Survivors* sits on a secure cloud server, hosted by DFnet.

### Data and Statistical Analyses

#### Aim 1

Demographic and risk factor data will be analyzed/reported using descriptive statistics. Bivariate analyses will be employed to examine key clinical and laboratory test differences between EVD survivors and controls using chi-square and two sample t-tests as needed. Unadjusted estimates of the proportion of patients with uveitis in the two groups will be calculated. Among those who develop uveitis in the cohort of EVD survivors and to determine independent predictors of uveitis, a multivariate logistic regression will be constructed with odds ratios and 95% confidence intervals of the odds ratios. This model will include pertinent clinical and demographic variables, not limited to age, gender, medical history, and ocular symptoms. Time from acute EVD will be included in the multivariable regression model as a potential confounding variable.

#### Aim 2

The Chi-square test or Fisher’s exact test will be used to compare the proportion of EVD survivors with detectable EBOV RNA by RT-qPCR of the vitreous fluid with the control subjects. The Jeffrey’s interval approach will be employed to calculate a 95% confidence interval to estimate the true proportion of EBOV RNA-positive vitreous specimens within the EVD survivor group [20]. Either the unpaired t-test or Mann-Whitney U test, as appropriate, will be used to compare median GWC values for EBOV-specific antibody between EVD survivors and the control subjects.

#### Aim 3

The relationship between uveitis and lymphocyte markers will be determined by bivariate analyses, as appropriate, in addition to an analysis to examine the effect of viral load at the time of acute EVD. Bivariate analyses will compare these to expression of effector, activation, and homing markers on T-cells and B-cells, along with Ig subtype using t-tests or correlation coefficients, as required. A multivariate linear regression model will be created and include initial viral load, ocular symptoms, age, gender, time from acute EVD, and other statistically relevant immune phenotype variables.

For each Aim, SAS® v9.4 will be utilized for statistical analyses and a p-value of 0.05 will be the threshold for statistical significance.

### Safety Considerations and Monitoring

Due to the potential for persistence of EBOV in intraocular tissues or fluids, appropriate personal protective equipment (PPE) and IPC practices will be utilized for any invasive ophthalmic procedures. As part of the EVICT Study, we designed an ophthalmic procedure room with high-level infection control measures according to guidance from Emory University and the World Health Organization (WHO) [20]. Standard Operating Procedures (SOPs) have recently been developed for vitreoretinal surgery, in partnership with experts in infectious disease, ophthalmology (including retina and uveitis expertise), public health, global health, nursing, and experts in biocontainment (Figure 2 and Supplemental Material).

**Figure 2.**
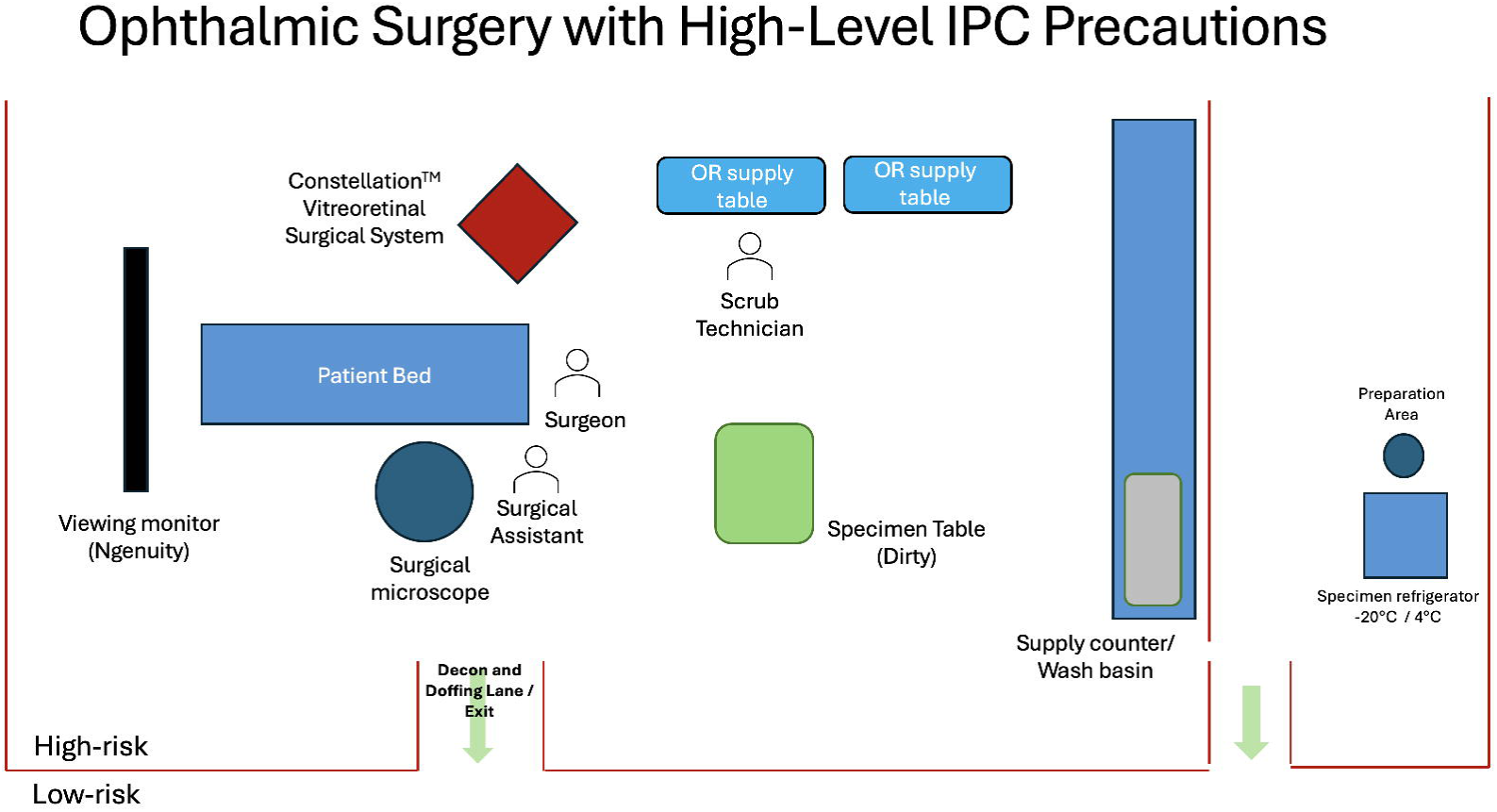
Schematic diagram of operating room for ophthalmic surgery targeting the pars plana vitrectomy procedure with maximum containment IPC precautions. Key features of the operating room design include: a clear space for doffing where one transitions from high-risk to low-risk areas; a specimen (dirty) table is available for placement of specimens; disinfection of primary and secondary specimen containers; and a hand sanitizing station for specimen packaging and transport to appropriate refrigeration. Strict disinfection protocols are implemented for instrument entry into and exit from the eye, as per the protocol outlined in the Supplemental Material.

Study investigators from the United States and Sierra Leone will review data collected from this study. Enrolled participants will be monitored for complications for up to 6 months. If an increase in complications among study participants occurs, the study procedures and associated safety protocols will be reevaluated with collaborators to minimize increased risk. The University of Nebraska Medical Center IRB and the Sierra Leone Ministry of Health and Sanitation Office of Ethics will be notified in the case of any reportable events.

## DISCUSSION

In the aftermath of the 2013-2016 Western African EVD outbreak and multiple subsequent outbreaks in DRC, thousands of EVD survivors are at risk of serious post-acute sequelae requiring ongoing individual evaluation and care and that may have public health implications. These protocols outlined in this study bridge clinical characterization and the investigation of the molecular virologic and immunologic features of EVD-associated uveitis. Specifically, the molecular assessment of EBOV in ocular tissues and fluids, paired with characterization of local and systemic EBOV-specific immune responses, may delineate aspects of pathogenesis and identify novel immunologic biomarkers with which to stratify risk and disease severity and target preventive or therapeutic interventions. Any relationship between viral persistence and immunologic biomarkers in patients with EVD-associated uveitis may inform understanding of similar phenomena in other FVDs as well as other infectious diseases in which viral persistence is associated with uveitis (e.g., in the setting of rubella virus [25], Zika virus [26], and HIV infections [27, 28])

Given the long-standing, multi-disciplinary partnerships with academic institutions, non-governmental organizations, and local government entities, we aim to implement this program to inform future ophthalmic care for EVD survivors and contribute to the growing body of knowledge pertaining to EVD survivorship in the anticipated event of future outbreaks. Additionally, the clinical research infrastructure developed in partnership with Sierra Leone government and non-government stakeholders will provide critical vision health, educational, and research infrastructure to address vision health disparities in Western Africa beyond EVD survivor care.

Continued engagement with EVD survivor advocacy groups and the Sierra Leone MOH will promote participation from those impacted by EVD, as well as communication for the communities, countries, and regions impacted by the threat of new outbreaks. Given the ongoing threat of EVD (and other FVD) re-emergence in Western African countries as well as in Central Africa in recent years, this study will provide critical information regarding long-term sequelae in the post-FVD setting, and operational and preparedness measures that could be deployed to other countries in FVD-affected countries in the future.

At the conclusion of the study, pertinent findings will be disseminated to local communities and healthcare stakeholders, local government officials, and the broader medical community through presentations, health reports, and journal publications. These communications will inform preparedness measures for EVD and other FVD and guide survivor care programmes, and could also inform other, emerging infectious diseases, including other WHO high impact pathogens that may also present a risk to vision health [29].

## Data Availability

All data produced in the present study are available upon reasonable request to the authors

## Acknowledgements

We thank those that enrolled in the EVICT Study and their family and friends who supported them throughout their involvement in this work. We would also like to thank the Sierra Leone Ministry of Health and Sanitation National Eye Programme, Sierra Leone Association of Ebola Survivors, Central Global Vision Fund, the Lowell and Ruth Gess Eye Hospital, Connaught Government Hospital staff, and the Kissy United Methodist Church Medical Board.

## Funding and Acknowledgements

This project was supported by the National Eye Institute of the National Institutes of Health under award number R01 EY029594 (Steven Yeh) and K23 EY030158 (Jessica Shantha). Work on this project has also been partially funded in part with federal funds from the National Cancer Institute, National Institutes of Health, under Contract No. 75N91019D00024 (Ian Crozier). The content is solely the responsibility of the authors and does not necessarily reflect the views or policies of the Department of Health and Human Services, nor does mention of trade names, commercial products, or organizations imply endorsement by the U.S. Government.. Funding support is also provided by the Macula Society, the Retina Society, Association for Research in Vision and Ophthalmology, and the Stanley M. Truhlsen Family Foundation, Inc. The opinions, interpretations, conclusions, and recommendations presented are those of the author and are not necessarily endorsed by the U.S. Army.

## Notes

### Competing Interest Statement

The authors have declared no competing interest.

### Author Declarations

Ethics committee/IRB of University of Nebraska Medical Center gave ethical approval for this work. Ethics committee/IRB of Emory University School of Medicine gave ethical approval for this work. The Ministry of Health Office of Ethics Approval in Sierra Leone gave ethical approval for this work.

