## Supplemental 1 for "Understanding the pathogenesis of uveitis in Ebola virus disease survivors: a study protocol for clinical, molecular virologic, and immunologic characterization"

**Supplementary File**

**Sierra Leone Ministry of Health and Lowell and**

**Ruth Gess Eye Hospital Ebola Survivor (SMILE)**

**Study Group Roster**

**University of Nebraska Medical Center,** *Omaha, NE, USA*

Steven Yeh, MD, Christopher D. Conrady, MD, PhD, Caleb Hartley, MPH, Tolulope Fashina, MD, MPH, Angela Hewlett, MD, MS, David Brett-Major, MD, MPH, Michael Wiley, PhD, Ye Huang, MD, Charlene Choo, MD, Brent R. Hayek, MD, Helen Song, MD, Ileana Fortune, Lucas Kim, MD, Jessica Gard, Shiama Balendra, Caleb Yeh, Crystal Huang

**Ministry of Health National Eye Programme,** *Freetown, Sierra Leone*

Lloyd C. Harrison-Williams, MD, John G. Mattia, MD, Jalikatu Mustapha, MD, Matthew J. Vandy, MD

**Ministry of Health,** *Freetown, Sierra Leone*

Zikan Koroma, MD, Alie Wurie, MD, Theophilus Frankly, Osman Conteh, Benjay, Alicious Kamara, Santigie

**Lowell and Ruth Gess Eye Hospital,** *Freetown, Sierra Leone*

Ibrahim Conteh, Agnes Konneh, Ken Campbell, Mabel Nanah Sesay, Miatta Dean Rogers, Rosamond Hannah Dowrie, Ramatu Lahai, Mohamed Mansaray, Philip Koroma, Mohamed Kamara, Ibrahim Cotor Bah, Tamba George Mani, Benjamin Williams, Dr. Lloyd Harrison-Wiliams

**Kenema Government Hospital Lassa Hemorrhagic Fever Laboratory,** *Kenema, Sierra Leone*

Augustine Goba, John D. Sandi, Mambu Momoh, Simbirie Jalloh, Donald S. Grant, MD, MPH

**Clinical Monitoring Research Program Directorate, Frederick National Laboratory for Cancer Research,** *Frederick, MD, USA*

Ian Crozier, MD

**Emory University,** *Atlanta, GA, USA*

Jessica Carag, DVM, Jill Morgan, RN, BSN, Colleen S. Kraft, MD, MSc, Susanne Linderman, PhD, Rafi Ahmed, PhD, Laura Ward, MPH, Sanjana Kuthyar, MD

**University of California San Francisco,** *San Francisco, CA, USA*

Nisha Acharya, MD, MS, Thuy Doan, MD, PhD, Jessica G. Shantha, MD, MSc

**Flinders University,** *Adelaide, Australia*

Justine Smith, PhD

**Tulane University School of Medicine,** *New Orleans, LA, USA*

John S. Schieffelin, MD, Robert F. Garry, PhD

**George Mason University,** *Fairfax, VA, USA*

Carolyn Drews-Botsch, PhD, MPH

**United States Centers for Disease Control and Prevention,** *Atlanta, GA, USA*

Timothy M. Uyeki, MD

**University of North Carolina-Chapel Hill,** *Chapel Hill, NC, USA*

Jean-Claude Mwanza, MD, PhD, MPH

**Sierra Leone Association of Ebola Survivors,** *Freetown, Sierra Leone*

Mohamed Mansaray, Yusuf Kabba, Daddy Kamara

**United States Army Medical Research Institute of Infectious Diseases,** *Ft. Detrick, MD, USA*

Xiankun (Kevin) Zeng, PhD

**Central Global Vision Fund,** *Millbank, SD, USA*

Roger Reiners, Melanie Reiners

**World Health Organization,** *Geneva, Switzerland,*

Anaïs Legand, MPH, Pierre Formenty, DVM, MPH

**Walter Reed National Army Medical Center,** *Washington, D.C., USA*

Grant A. Justin, MD
