## Supplemental 3 for "Understanding the pathogenesis of uveitis in Ebola virus disease survivors: a study protocol for clinical, molecular virologic, and immunologic characterization"

**Standard Operating Procedure – Vitreoretinal Surgery**

**Background**

This Standard Operating Procedure (SOP) document serves to highlight areas and practice patterns for infection prevention and control during vitreoretinal surgery for Ebola virus disease (EVD) survivors. The EVICT study demonstrated that at 18 and 34-month follow-up, Ebola virus RNA persistence was not identified in the intraocular fluid specimens in 50 patients anticipating cataract surgery or other ophthalmic surgery.^1^ This included 49 aqueous humor samples and 1 vitreous humor sample. The PREVAIL VII Study similarly showed that 22 EVD survivors anticipating cataract surgery tested negative for Ebola virus RNA in their aqueous humor.^2^ EVD survivors, however, remain at risk of uveitis that can lead to ocular complications within the posterior segment of the eye that require vitreoretinal surgery.^3^ Given the risk of Ebola viral persistence in intraocular fluids^4^ and tissues, including the vitreous,^5-6^ the SOP serves as preliminary guidance for invasive operative procedures for vitreoretinal surgery.

Patients who elect to have vitreoretinal surgery performed for a surgical indication (i.e., retinal detachment, vitreous opacity, vitreous hemorrhage) will be given the opportunity to participate in a clinical research study entitled “Pathogenesis of Uveitis in Ebola virus disease survivors”. For patients who undergo enrollment, vitreous humor will be collected for molecular virological, and immunologic analysis during ophthalmic surgical procedures performed at the Lowell and Ruth Gess Eye Hospital.

**Goals and Objectives**

The goals of vitreoretinal surgery in this context include:

1) Provision of safe and effective surgical care for the Ebola virus disease survivor with an indication for vitreoretinal surgery to improve or preserve visual function.
2) Mitigation of the risk of peri-operative viral transmission for the patient, operative team, and surgeon, given the uncertain risk associated with vitreous fluid at this time.
3) Collection of intraocular fluid(s) and tissue(s), including vitreous specimens, for molecular virological and immune analysis by in-country partners, supported by UNMC, Emory Vaccine Center investigators, and Ministry of Health and Sanitation Sierra Leone National Eye Programme and Laboratory Medicine collaborators.

Considerations for this SOP include supply management, operative procedure, management of instrumentation during the surgical procedure, specimen management and waste management.

**Supplies**

The supplies needed include vitreoretinal equipment and cassettes, PPE for the surgical scrub tech, OR staff, and retina surgeon, intraocular instrumentation (i.e., laser, soft tip cannula, membrane forceps), syringes (3-, 5-, and 10-cc syringes; RNA later) as required for diagnostic analysis purposes.

Infrastructural Requirements

Vitreoretinal Surgical System - Alcon Constellation® Vision System (Alcon, Ft. Worth, Texas, USA)

Gas with medical grade air or nitrogen and tubing

Gas regulator for PSI between 100-120 psi

Stable power supply via generator, Universal Power Supply (UPS) devices for microscope and VRS system

Vitrectomy supplies include the following:

25-gauge vitrectomy pack including light pipe, vitrectomy handpiece, infusion cannula, trocar cannula system

25-gauge endolaser (Illuminated endolaser preferred)

3-way stopcock

Balance saline solution or lactated Ringers solution

1000 centistoke silicone oil or intraocular gases (SF6 or C3F8)

6-0 or 7-0 Vicryl suture

Scleral depressor

Syringes and specimen collection

3-cc syringe

5-cc syringe

10-cc syringe

25-gauge needle

Flocked or polyester swab with universal or viral transport media

PPE

Surgical gowns

Surgical gloves

Eye protection

Surgical mask

Sterilization

Basin

5% Povidone iodine

Sterile H20

Sterile 4x4s (2-3 packs per patient)

Cotton swabs (2-3 packs per patient)

Retrobular block supplies

25-gauge 1 ½” needle

2% lidocaine without epinephrine

0.75% Marcaine 50:50 mixture

Alcohol swab

4x4 swabs

S, M, L and XL gloves

Sticker or marker to designate operative eye

Medical waste handling:

Sharps containers

Biohazard bags and bins

Non-biohazard bags and bins

**Protocols and Procedures**

Preoperative

*Universal triage step*

1. Verify that the patient undergoing the vitreoretinal surgery and indicated ophthalmic procedures is free of symptoms as well as communicable risk.
2. These symptoms include fever greater than 100.4° Fahrenheit (38° Celsius), active diarrhea, vomiting, cough, or shortness of breath.
3. Obtain additional workup as needed for any of the above symptoms prior to proceeding with surgery.

*Retrobulbar block*

1. Patient’s eye is marked, and consent verified.
2. Alcohol applied to inferior periorbital rim.
3. Retrobulbar block administered x 7-8 cc in standard fashion at 1/3 from lateral canthus for proptosis.
4. Manual pressure to decompress the globe.
5. Patient waits 8-10 minutes prior to entering operative environment (if performed in anteroom).

Operative Protocols

1. Operative staff don appropriate PPE for ophthalmic surgical protocol and under direct observation.
2. Surgical scrub prepares vitreoretinal surgical system and surgical settings as per routine.
3. Additional alcohol-based hand sanitizer available on neighboring table to be utilized at the time of specimen collection.
4. Time out taken to verify patient, name, surgical site, procedure to be performed.
5. Patient prepped, draped in sterile fashion.
6. Surgeon and scrub tech scrub and sterile for procedure.
7. Addition small table of ‘extras’ that have been inside the eye (Dirty table)
8. 25-gauge cannulas introduce by surgeon; Trocar placed on Dirty table.
9. Diagnostic PPV initiated with 3-way stopcock and assistant withdrawing specimen (aspirating from syringe)
10. Surgeon unhooks 3-cc syringe from 3-way stopcock and places specimen on Dirty Table. Surgeon uses alcohol-based hand sanitizer to foam hands.
11. Surgeon attaches 5-cc syringe to 3-way stopcock and performs vitrectomy with assistant aspirating fluid with the infusion on.
12. Surgeon unhooks 5-cc syringe from 3-way stopcock and places specimen on Dirty Table. Surgeon uses alcohol-based hand sanitizer to foam hands.
13. Surgeon attaches 10-cc syringe to 3-way stopcock and performs vitrectomy with assistant aspirating fluid with the infusion on.
14. Surgeon unhooks 10-cc syringe from 3-way stopcock and places specimen on Dirty Table. Surgeon uses alcohol-based hand sanitizer to foam hands.
15. Surgeon performs vitrectomy including core and peripheral vitrectomy with scleral depression where needed.
16. Entry/exit of instruments
    a. When instrumentation is introduced, entry is as per routine. For exit, disposable instruments or reusable instruments are placed in the alcohol basin on the Dirty Table.
    b. When instrumentation is passed onto another Dirty Table (e.g., endolaser placed onto table connected to vitreoretinal surgical system), alcohol-based hand sanitizer should be used by the surgeon if the surgeon touches any part of the instrument that has gone into the eye.
    c. Otherwise, instruments connected to the machine should also be wiped down with 70% alcohol, laid on the dirty table and alcohol-based sanitizer used to foam the gloves of the surgeon.
17. Following complete vitrectomy and removal of all membranes (anterior-posterior and tractional forces), air-fluid exchange with the soft tip is performed to reattach the retina.
18. The soft tip cannula is removed and wiped down with alcohol. Alcohol-based sanitizer used to foam the surgeon’s hands and scrub tech where needed.
19. Silicone oil is instilled, and the 25-gauge injection catheter placed in the alcohol basin. Alcohol-based sanitizer used to foam the surgeon’s hands if the catheter is touched.
20. 7-0 or 6-0 vicryl suture used to close the sclerotomies to avoid oil / intraocular content extrusion.
21. Subconjunctival cefazolin and dexamethasone instilled.
22. A conjunctival swab is harvested and deposited in universal transport media or viral transport media.
23. Antibiotic ophthalmic ointment (e.g., erythromycin or equivalent) is instilled along with application of an eye patch and eye shield. Alcohol-based sanitizer used to foam hands.
24. Patient escorted to postoperative recovery area.

Specimen Handling

1. Sanitize gloves.
2. Primary containers for the specimens are wiped down with a 70% EtOH wipe, labeled, and dropped into a specimen bag (pre-labeled) from a receiver with appropriate PPE.
3. This bag should be dropped into a secondary container that is prelabeled.
4. Both the assistant and the surgeon (still sterile) should sanitize gloves and hands.
5. The specimen is placed in a 4-degree refrigerator prior to transportation to the lab.

Medical Waste Management

1. Verify disposal of sharps / Disposed in standard fashion.
2. All instruments on Dirty Table should be alcohol sanitized prior to autoclave.
3. Other medical instrumentation that is not reused should be wrapped by an operator who is gloved and experienced in medical waste management. Waste will be dropped in a large red biohazard bag.
4. This will be disposed of via medical incinerator.
5. All medical waste management activities will be overseen by personnel experienced in Infection Prevention and Control precautions according to Ministry of Health and Sanitation guidance.^7^

Exit for operative and ancillary staff: Operative staff doff appropriate PPE per standard protocol and under direct observation.

Additional Safety Considerations

1. In the event of a needlestick injury, incidents will be reported to the lead clinician on-duty and administrator on duty.
2. Needlestick injury protocols will be followed according to Ministry of Health and Sanitation, Sierra Leone guidance.^7^
3. In the event of a perceived safety breach, the lead clinician on duty and administrator on duty will be contacted to determine the nature of the breach, risk to the individual or individuals involved in the breach, and appropriate risk mitigation steps.

**Drafted by EVICT-VR Study Team Members**

Version 1.0: 26 Sept 2023

Version 1.1: 9 Jan 2024
